# Codesign of digital health tools for suicide prevention: A scoping review

**DOI:** 10.1101/2023.04.11.23288415

**Authors:** Dianne Wepa, Martin Neale, Waseem Abo-Gazala, Sally Cusworth, Jae Hargan, Manoj Mistry, Jimmy Vaughan, Stephen Giles, Mehnaz Khan

## Abstract

The importance of codesigning digital health tools for suicide prevention has gained popularity since 2012. Promoted as cost-effective and innovative, digital health tools are widely used but seldom described or evaluated from a codesign lens. This scoping review provides an overview of the research and gaps in the delivery of codesigned digital health tools for suicide prevention. This review is phase two within a three-phase study. Phase one involved a scoping review protocol which informed this scoping review and the results will contribute to a proof-of-concept project to develop a digital tool for suicide prevention (phase three).

The search strategy followed the Preferred Reporting Items for Systematic Reviews and Meta-Analyses Extension for Scoping Reviews (PRISMA-SCR) and Population, Concept, Context (PCC) framework to ensure reporting standards were maintained and supplemented by Arksey and O’Malley and Levac et al. The search dates occurred from November 2022 to March 2023. Five data bases were searched: Medline, Scopus, CINAHL, PsycInfo and Cochrane Database of Systematic Reviews. Grey literature searches included government, non-government health websites, Google and Google Scholar.

3260 records were identified from the initial search and 61 were included in the final review. All members of the research team screened the included records. Data from published and grey literature were extracted and a narrative approach identified the results and five themes (acceptability by users, future inclusion of experts-by-experience, inconsistent use of Patient and Public Involvement (PPI), digital tools to supplement face-to-face therapy and digital divide).

We found that none of the data from the included studies used codesign methodology and experts-by-experience roles were minimised as members of focus groups, advisory groups, pilot studies or at the final stage of usability testing. Future research is required where codesign involves co-authorship with experts-by experience, end-to-end partnership from design, implementation and evaluation of digital health tools for suicide prevention.

**Author summary:** As more people turn to digital technology (such as mobile apps and websites) to help with their mental health, they enjoy many of the benefits such as feeling less judged and being more affordable than face-to-face therapy. There are also risks involved such as how privacy is managed and reliance on the distressed person to self-manage their signs and symptoms. We found that people who have experienced suicidal thoughts and carers did not have an equal voice with those that developed the digital tools for suicide prevention. Our group comprising of experts-by-experience, health professionals, a mental health nursing student, technology expert and researchers felt that there was a gap in this area and met on a monthly basis for one year to see what the literature was saying. We found that the term codesign was used a lot but when we looked deeper into the articles and websites, we noted that experts-by-experience were only included to test apps or were involved in focus or advisory groups. We will be using the information from this scoping review to apply for funding to develop a digital solution that is truly designed with and by the people that need it the most.

## Introduction

Suicide affects more than 700,000 globally each year [1] and the ripple effect extends to the person’s family, friends and community [2]. As a response to prevent suicide, over 10,000 digital health technologies (video conferencing, smartphone apps, texting and social media) emerged as the first port-of-call for people experiencing mental health distress especially during and immediately after the COVID-19 pandemic [3–7]. Given the exponential rise of digital solutions for suicide prevention, there appears however, to be a disconnect between bringing the technology to market and involving experts-by-experience or service users as codesigners [8]. With a rapidly evolving technological environment, understanding the place and space of experts-by-experience in suicide prevention requires urgent attention.

For this scoping review the following definitions of codesign and experts-by-experience are provided. Codesign is defined as ‘a co-creation approach involving collaboration between researchers and end users from the onset’ [9] (p. 1). Experts-by-experience is a term that was selected by our research Patient and Public Involvement (PPI) group members, as it reflected the range of experiences as carers and mental health services users. Experts-by-experience is therefore defined as ‘people who have recent personal experience of using and caring for someone who uses health, mental health and/or social care services ‘ [10].

This scoping review is phase two within a three-phase study. During phase one, our scoping review protocol outlined the process for the study which informed this review [11]. Results from this scoping review will inform a funding application to codesign a proof-of-concept digital health tool for suicide prevention (phase three).

## Materials and methods

We used the recommended six stages of scoping reviews [12,13] and the Preferred Reporting Items for Systemic Reviews and Meta-Analyses Extension for Scoping Reviews checklist (PRISMA-ScR) guidelines [14]. This review did not appraise the quality of evidence found as it sought to examine a topic which has not been systematically reviewed before. We used the Population-Concept-Context (PCC) framework to strengthen the importance of this methodology [15].

### Stage One: Identifying the research question

Our research question was: *What is known from the existing literature about codesign of digital health tools for suicide prevention*. The PPC framework assisted with guiding us to identify the important aspects of the research question and determining the inclusion and exclusion criteria. We conducted our search from November 2022 to March 2023. The population of interest included patients, students, adults, internet users, experts by experience, service users and soldiers. Concept included suicide intervention, reduction and prevention. Context were countries such as New Zealand, UK, USA, Australia and countries that produced English language as primary and secondary literature within the European Union. The search timeframe was from 2012-2023 to ensure recency of the evolving nature of digital technology. The exclusion criteria were study protocols and opinion/editorial data. The search strings for published and grey literature are provided in Table 1.

**Table 1.** Population, Concept, Context & Search String.

| Population | Search String |
| --- | --- |
| Patients<br>Adults<br>Internet users<br>Experts by experience<br>Students<br>Pupils<br>Soldiers<br>Service users | Patient* OR student* OR "internet user*" OR<br>Adult* OR pupil* OR soldier* OR "experts by experience" OR "service<br>user*" |
| Concept |  |
| Suicide intervention<br>Suicide reduction<br>Digital intervention<br>Suicide prevention | Softwar* OR app* OR tool* OR technolog* OR digital* AND suicide*<br>OR "suicide prevent*" OR "suicide reduc*" OR "suicide intervent*" OR<br>MH* OR "internet based" OR "web based" OR "mobile based" |
| Context |  |
| United Kingdom<br>Australia<br>United States of America<br>New Zealand<br>European Union<br>2012-2023 | >2012 AND PUBYEAR >2023<br>AND English lang*<br>UK* AND United King* OR US*<br>United State* AND Canad*<br>AND NZ* OR New Zeal*<br>AND European Un* OR EU* |

### Stage Two: Identifying relevant sources

#### Published literature: Search strategy

The following licensed electronic databases (from 2012 to 2023) were systematically searched: Medline, Scopus, CINAHL, PsycInfo and Cochrane Database of Systematic Reviews. The initial sources were limited to English in the UK, Canada, USA, Australia and New Zealand. After receiving feedback during the review stage of our protocol, we extended the context to include European Union countries with publications in English. Furthermore, we identified articles such as systematic reviews where the researchers were located in these respective countries, however the data they sourced were from countries such as China, Japan and Sri Lanka.

The initial published data of 3210 records were identified and uploaded into EndNote X9.0, a citation and management software. Almost half were duplicates and removed (n=1570) and the remaining 1640 articles were exported into the reviewing software, Covidence, for title and abstract screening before full-text screening (Fig 1) [16].

**Figure 1:**
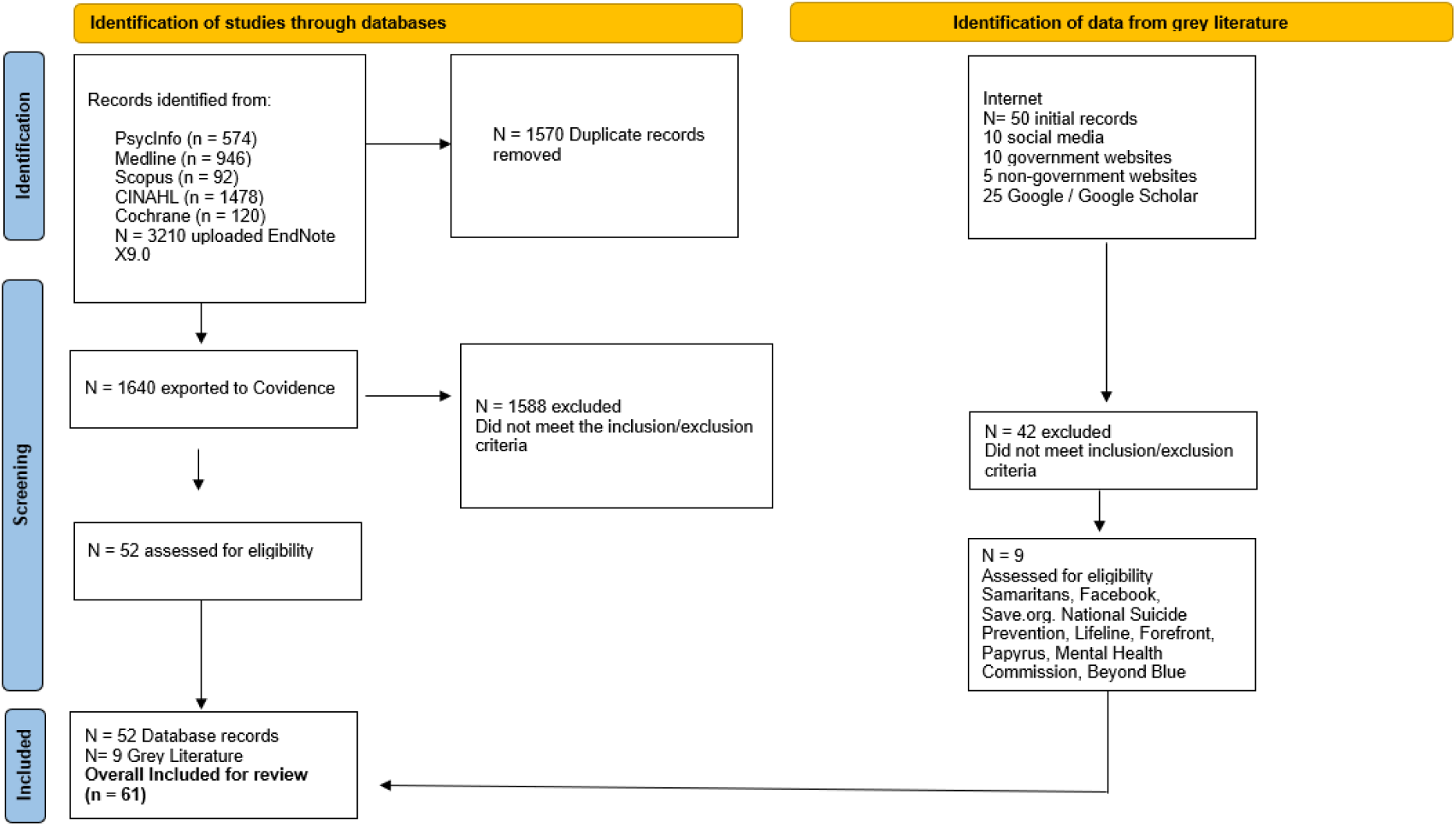
PRISMA flow diagram for included searches of databases and grey literature.

#### Grey literature: Search strategy

A grey literature search (from 2012 to 2023) was conducted using institutional and government electronic databases and websites. Google and Google Scholar were searched with relevant results reviewed which included non-government organisations and social media. The sources selected were limited to English. 50 records were identified with 42 excluded once the inclusion/exclusion criteria were applied and 9 records were assessed for eligibility. The included data were Samaritans, Facebook, Save.org, Lifeline, National Suicide Prevention, Forefront, Papyrus, Mental Health Commission and Beyond Blue (Fig 1).

### Stage Three: Source selection

From the-1640 articles uploaded into Covidence, the title and abstracts were screened by DW and WAB and MN resolved any conflicts. Full-text articles were discussed among the research team to ensure moderation of the process particularly with experts-by-experience. Once the inclusion and exclusion criteria were applied, we removed 1588 studies, leaving at total of 61 records (n=52 published articles, n=9 records from grey literature).

### Stage Four: Charting the data

All included data were first extracted from full-text articles and read and excerpts that were relevant to the research question were highlighted. Data from grey literature sources were also included in the data extraction table and included digital interventions predominantly from non-government websites. The data was charted according to the following categories: Author/Year, Aim/Objectives, Methods/Analysis, Population/Sample Size, Countries, Outcome/Limitations (Table 2).

**Table 2.**
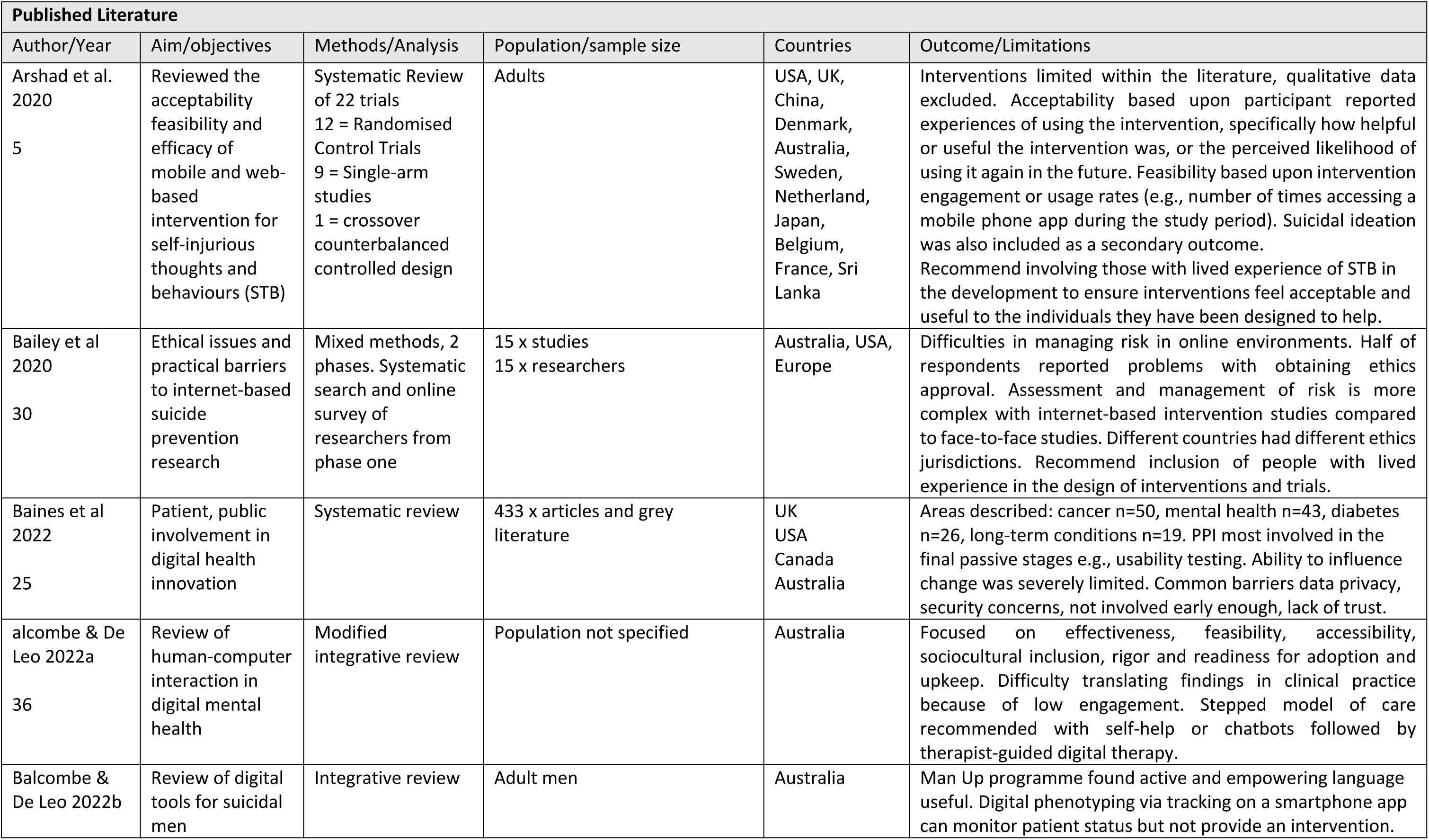
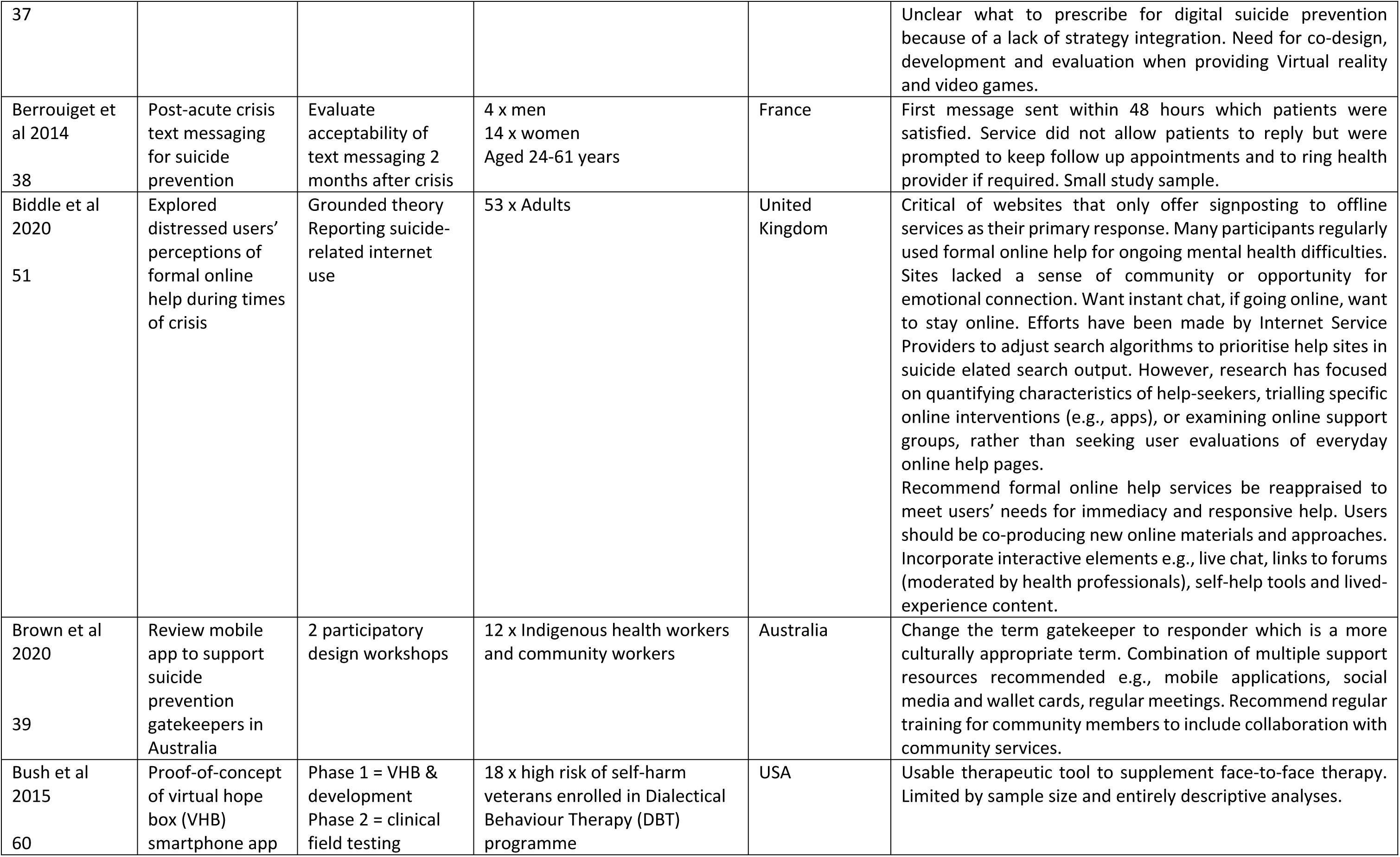
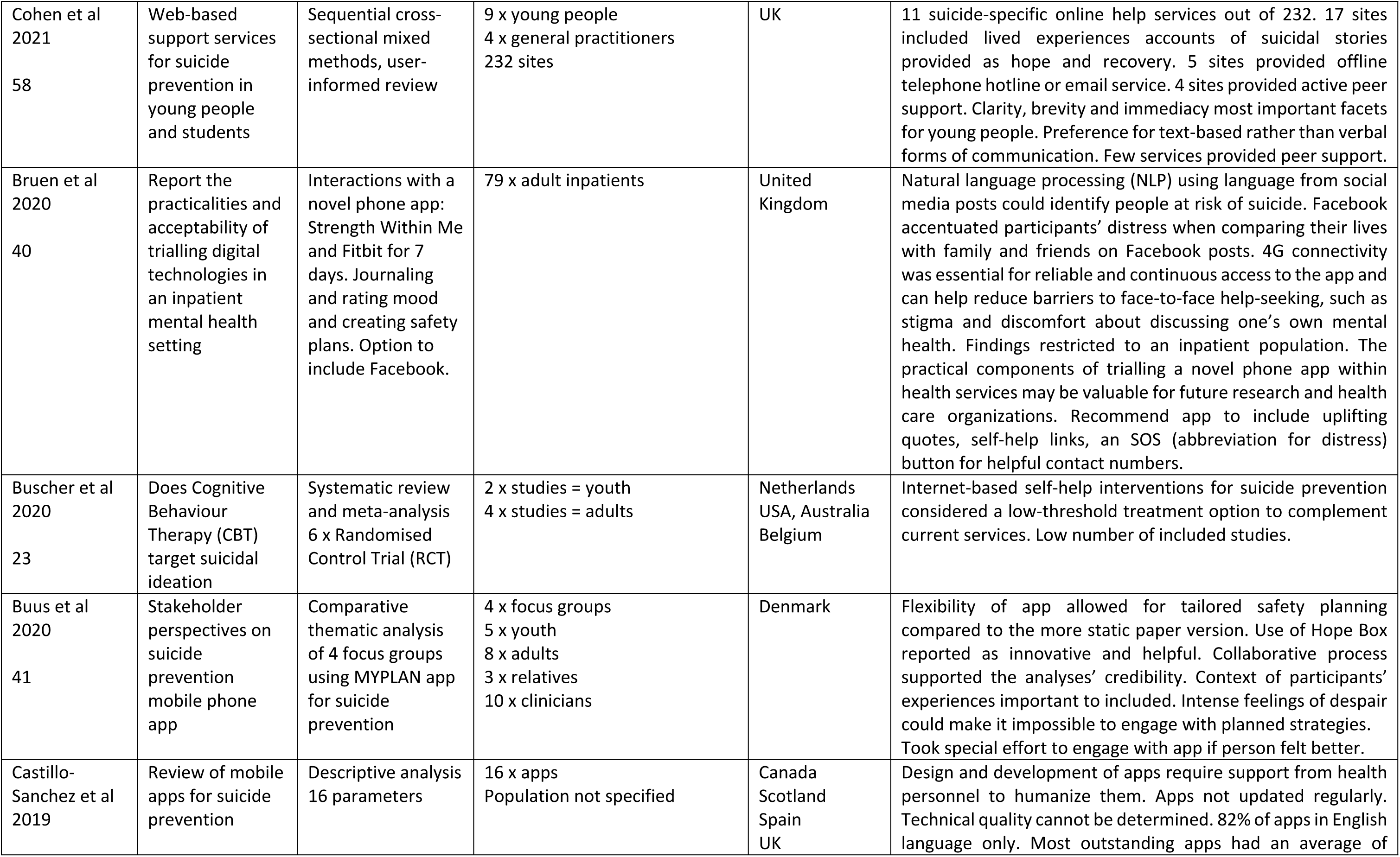
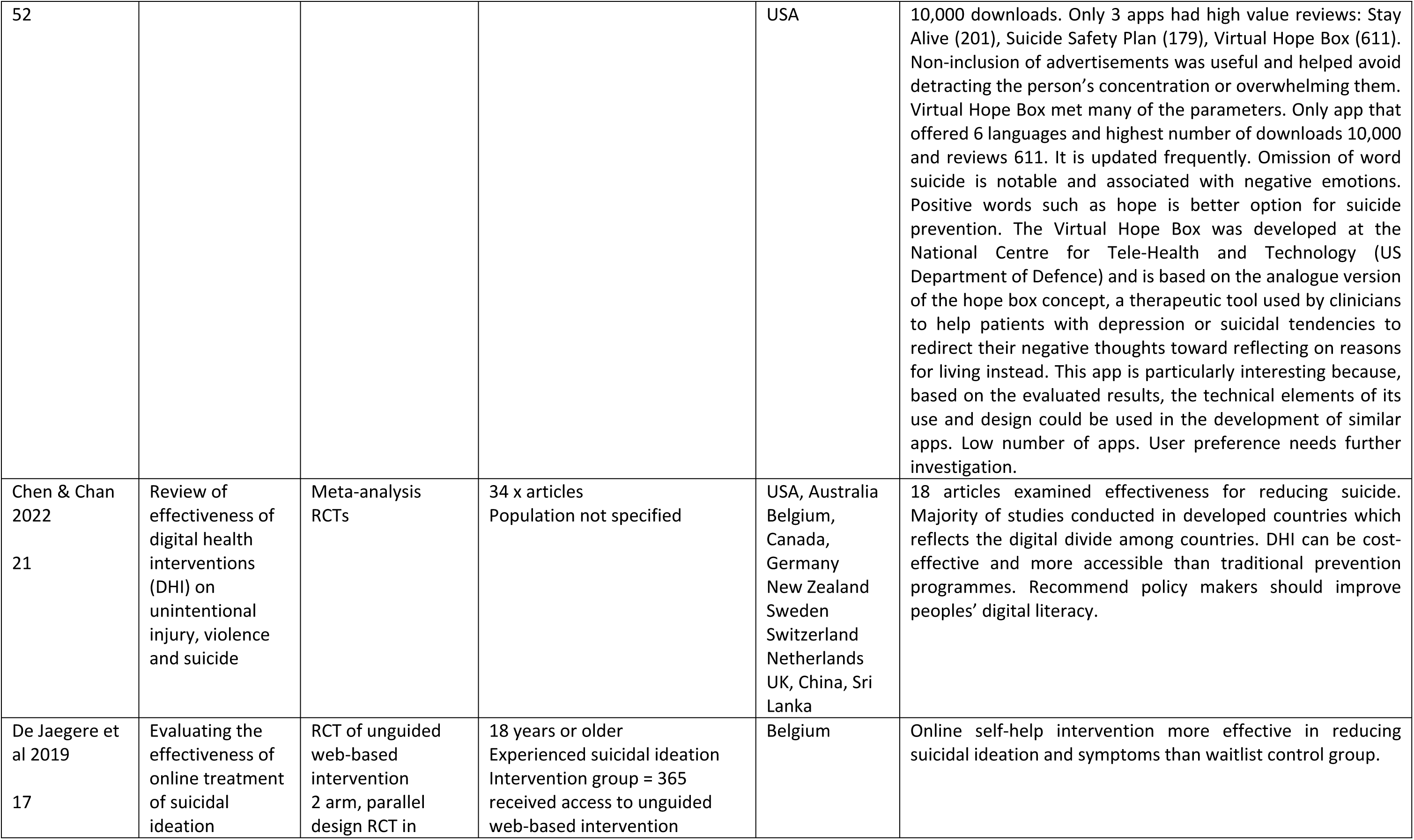
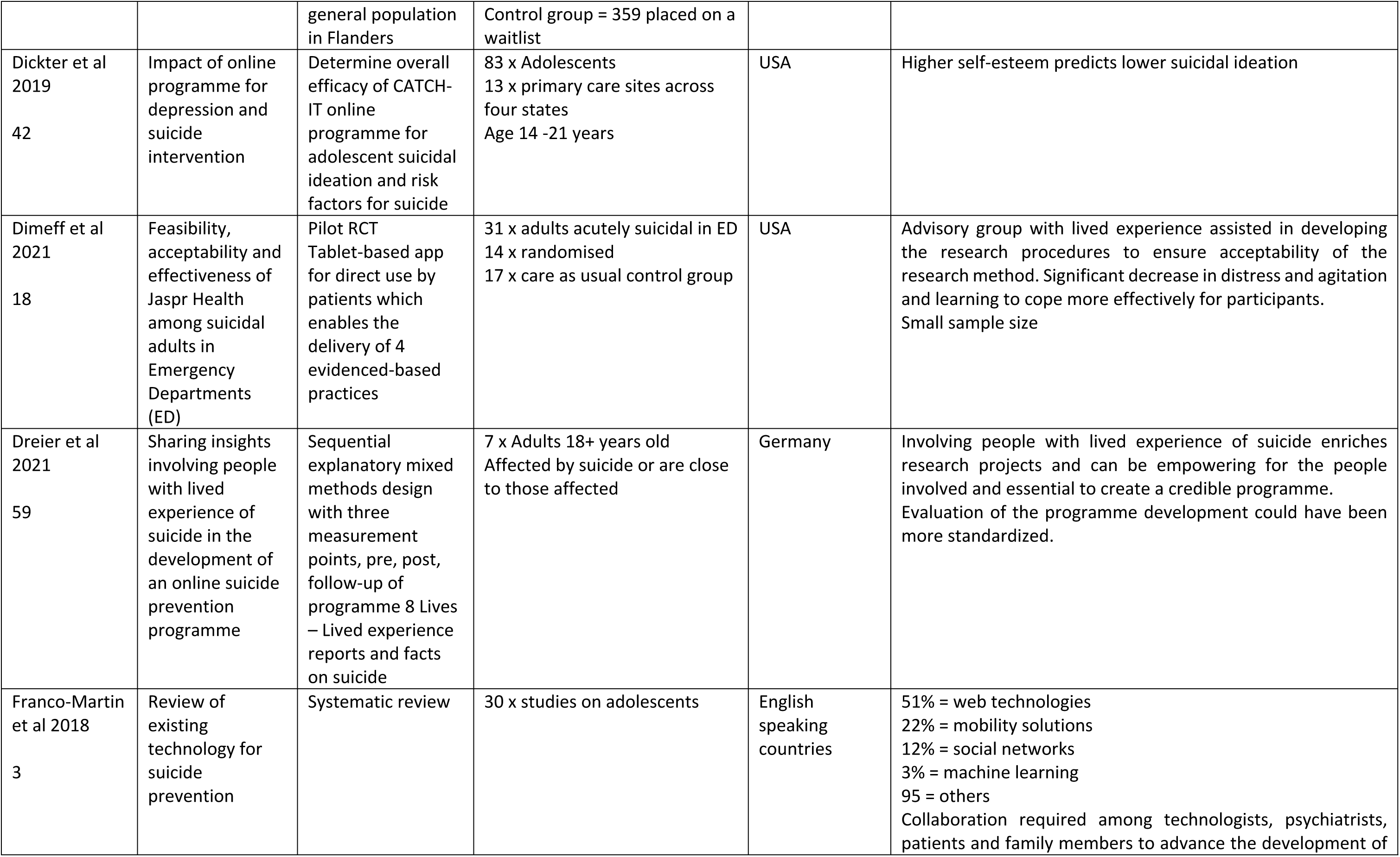
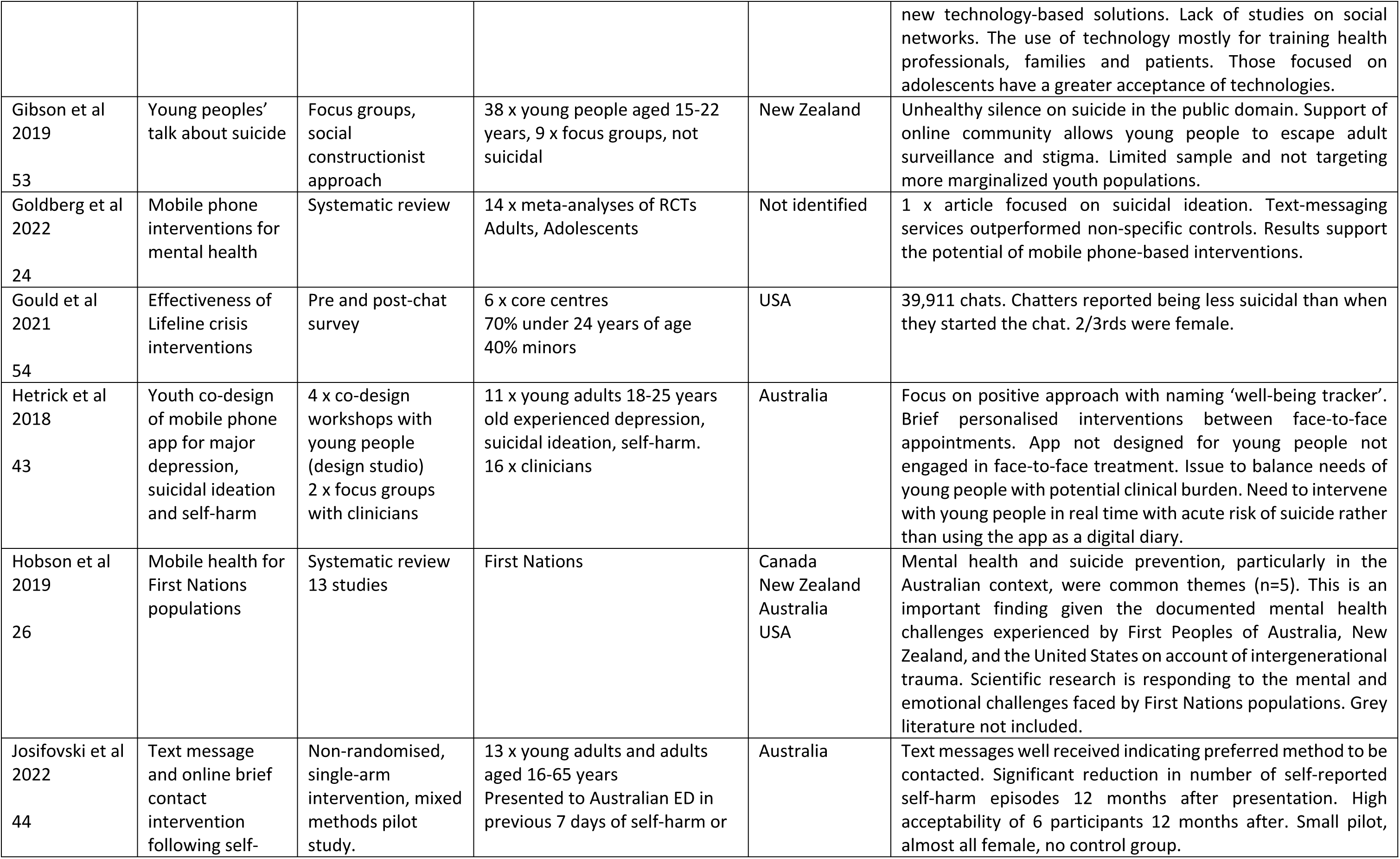
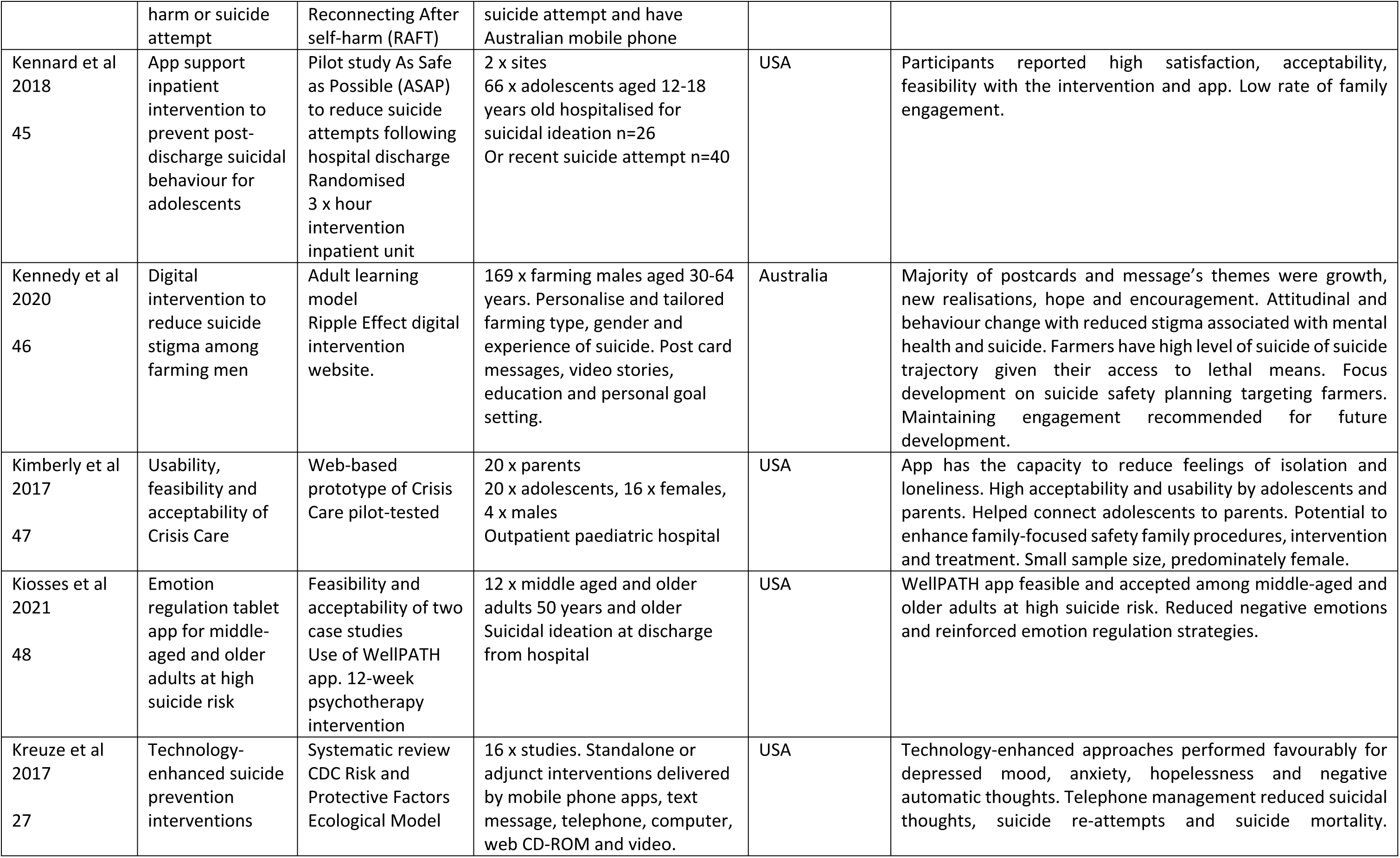
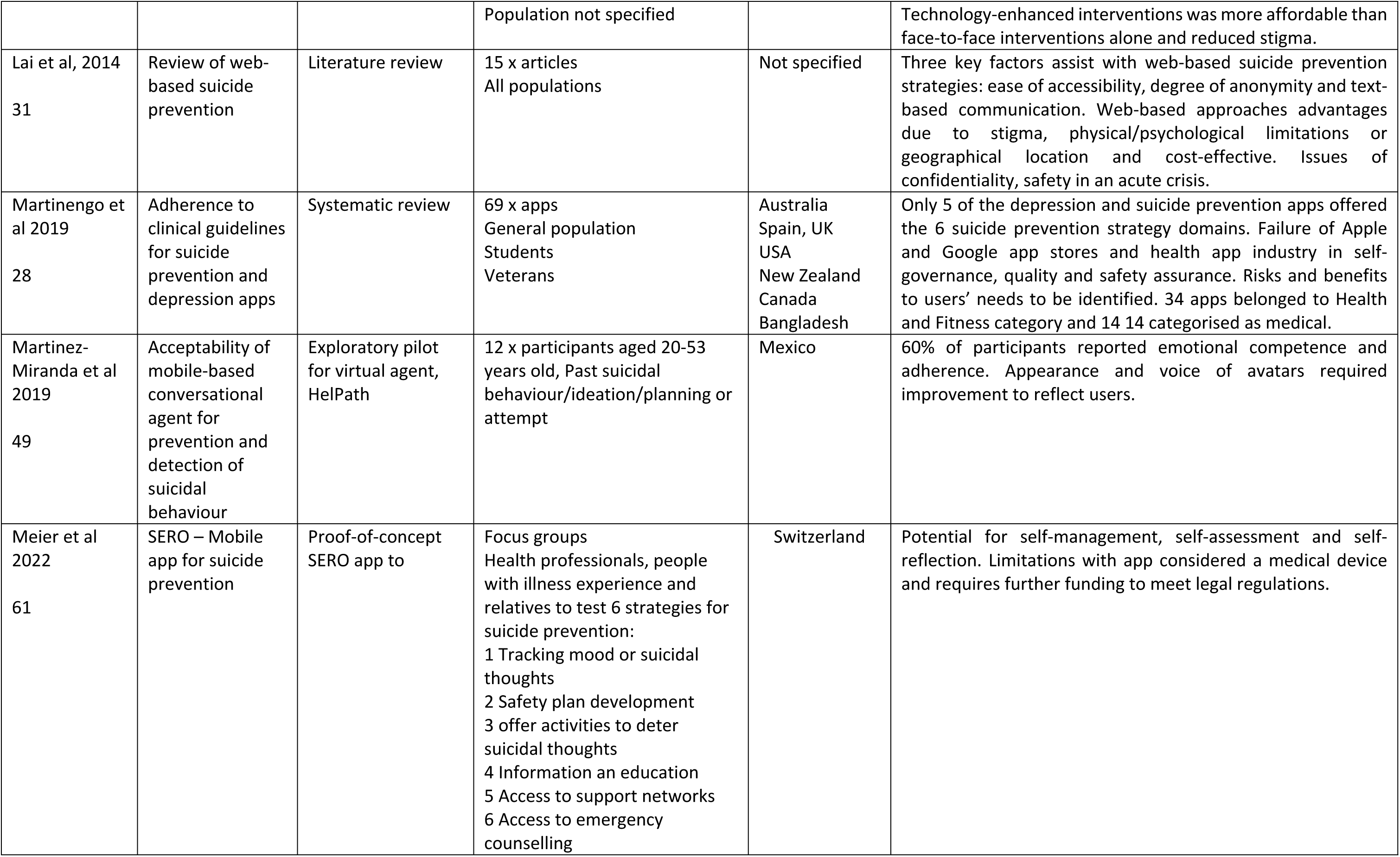
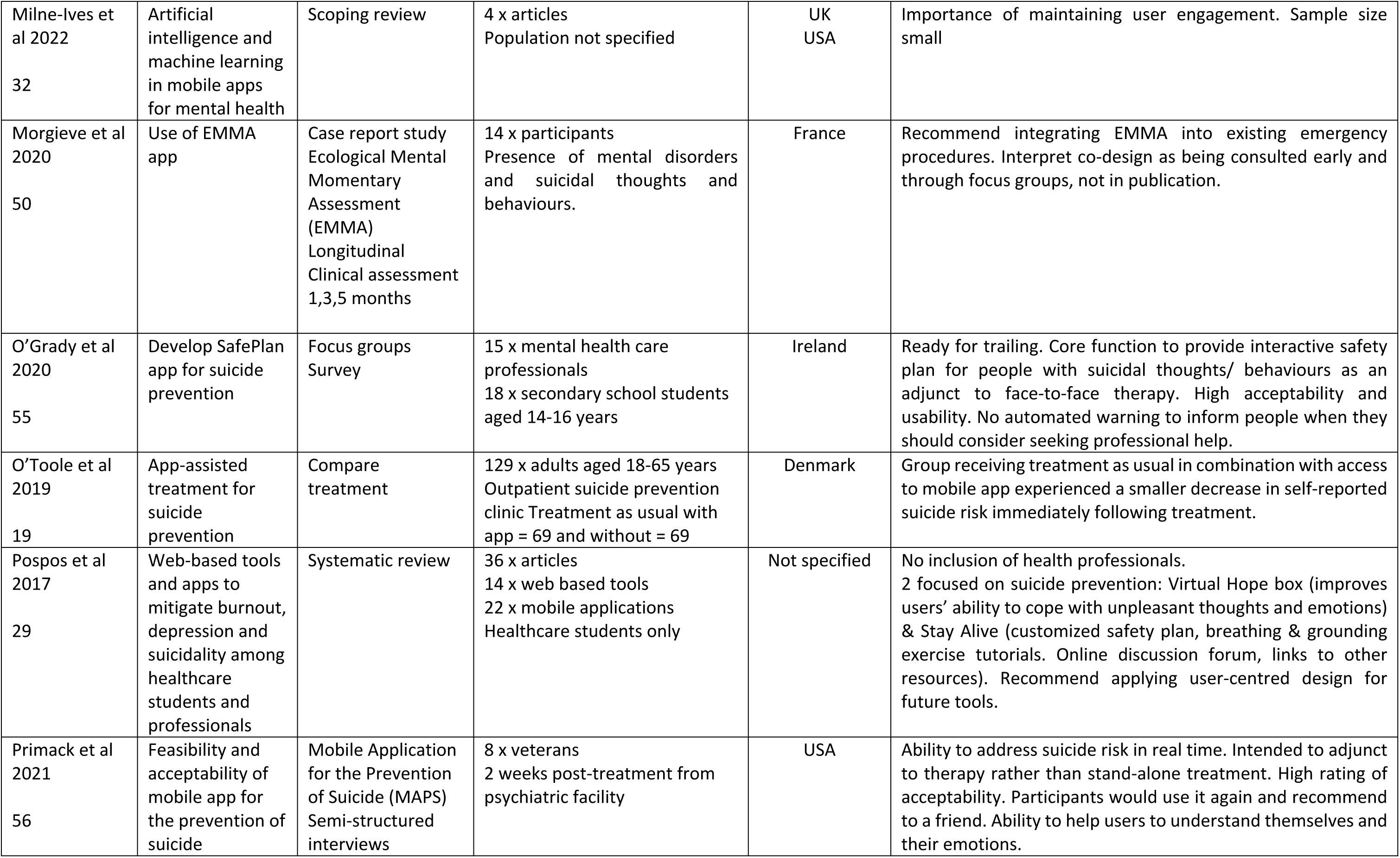
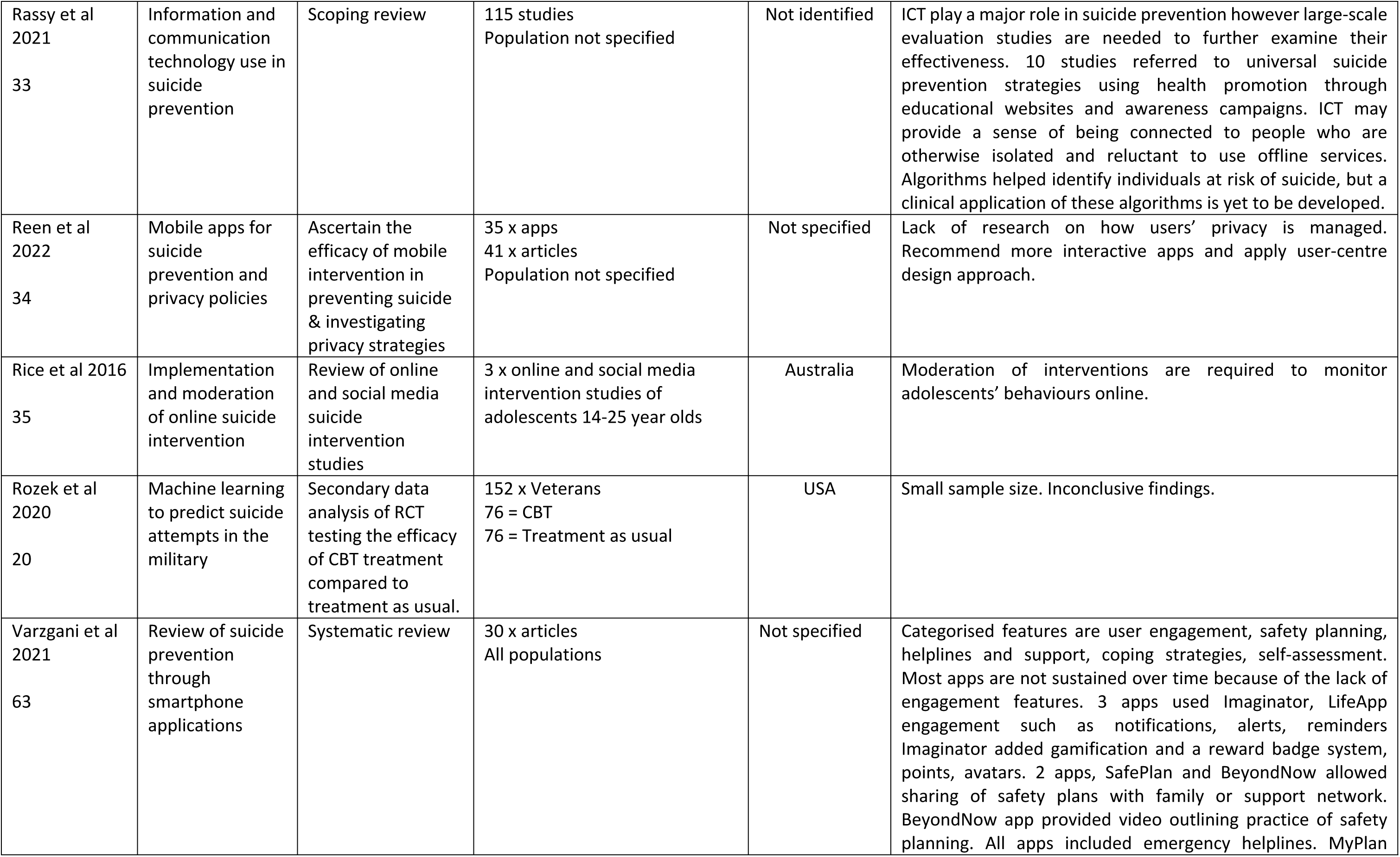
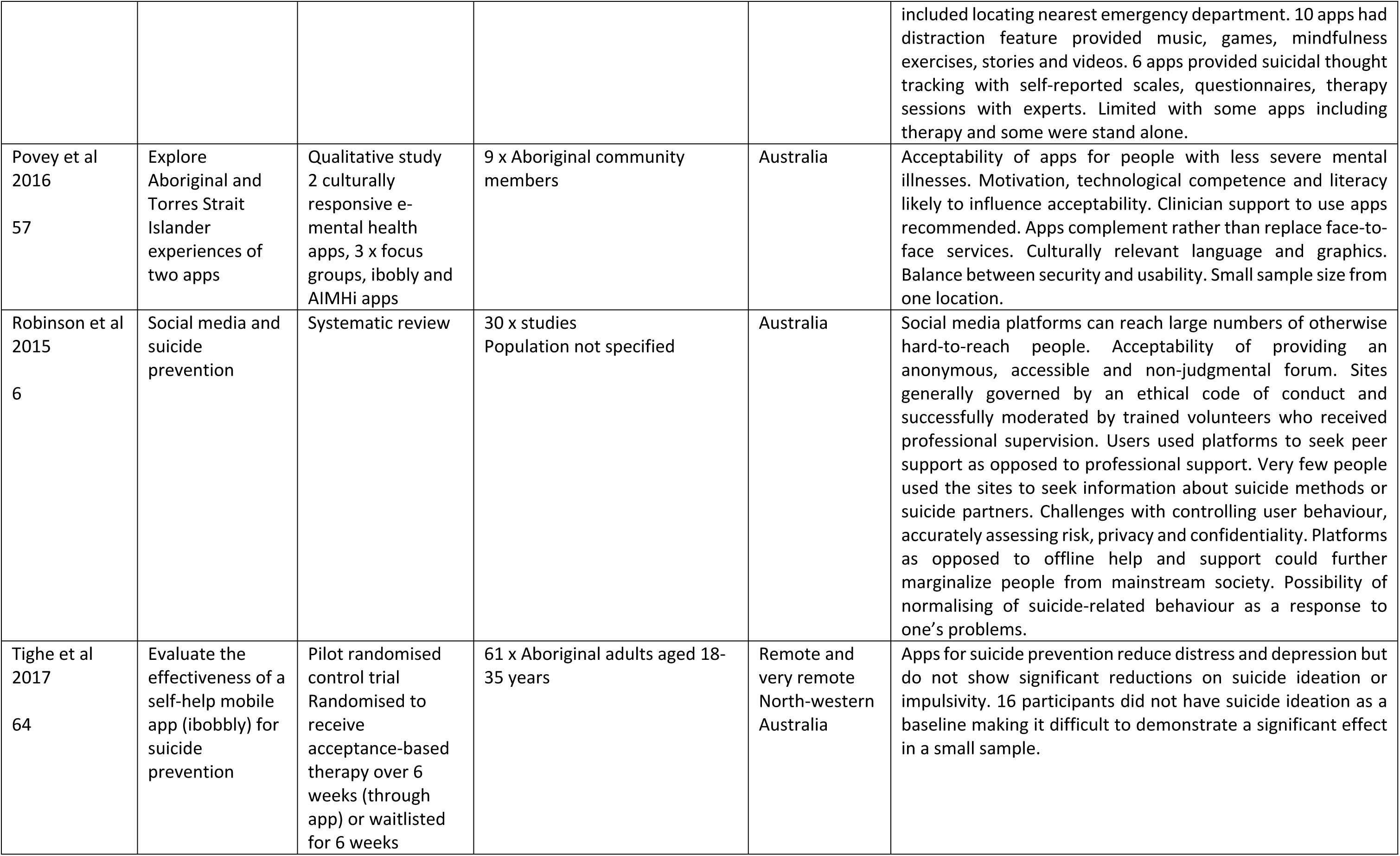
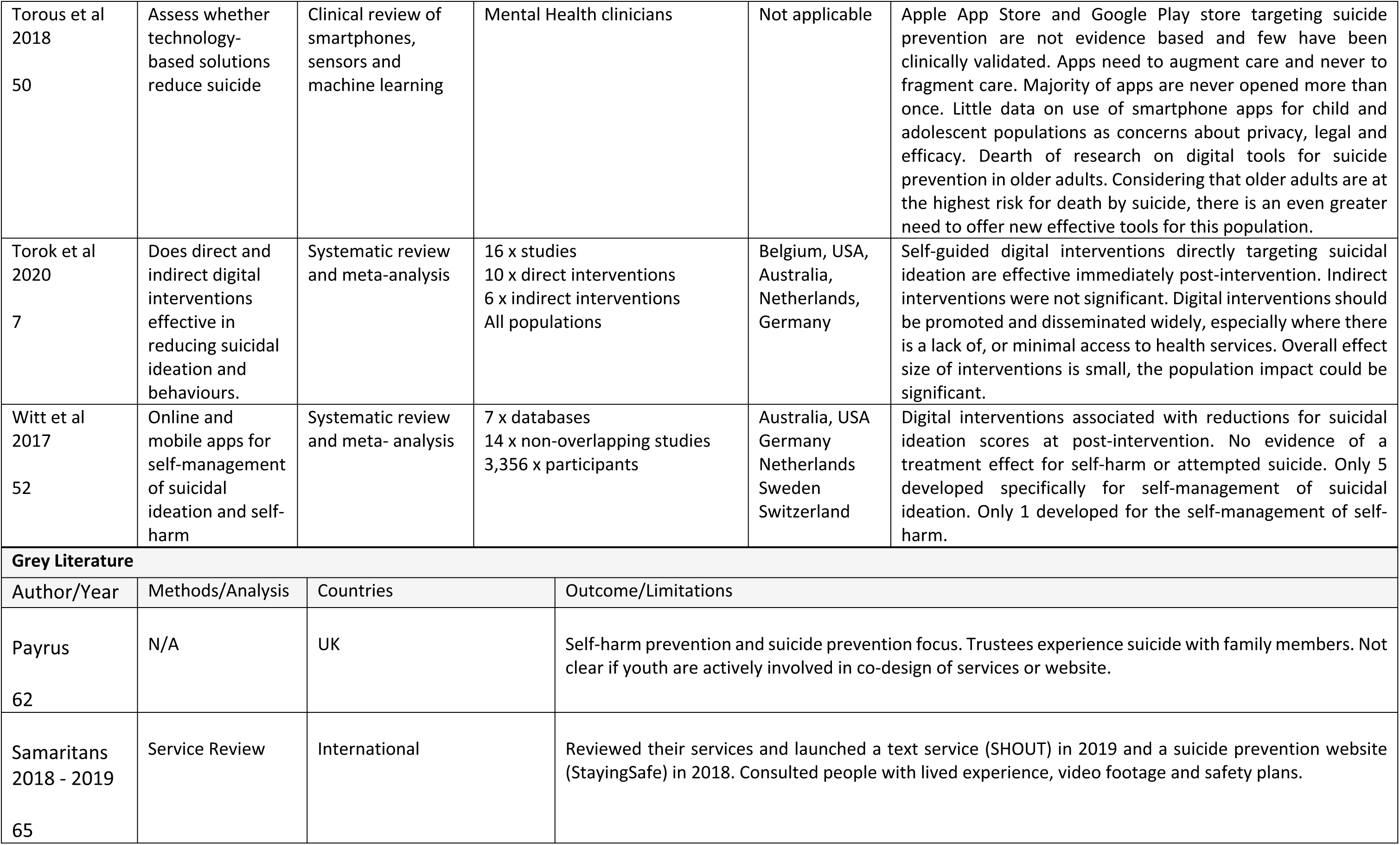
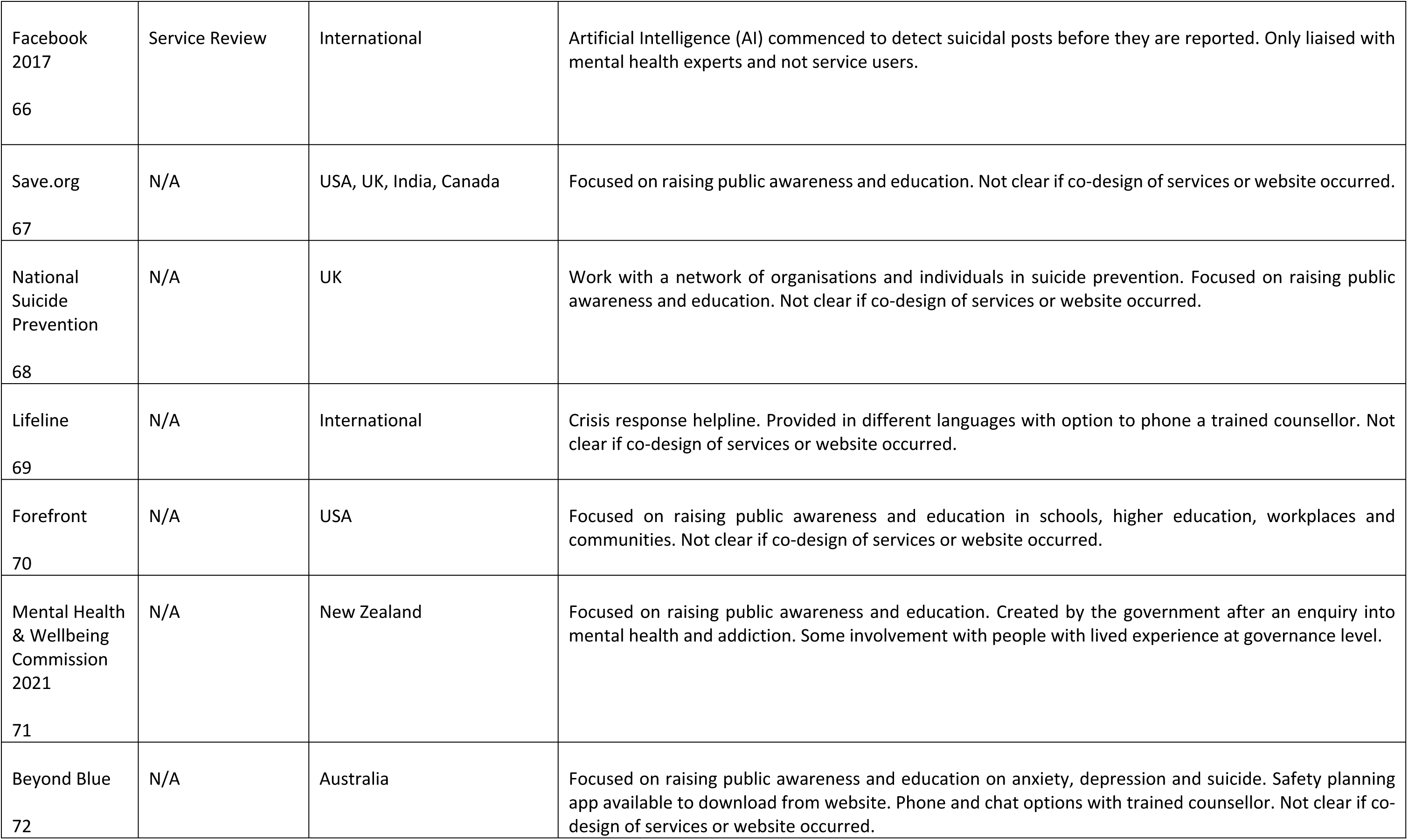
Data extraction table.

### Stage Five: Collating, summarizing and reporting the results

This scoping study aimed to present an overview of material reviewed which is not synthesised as is the case with a systematic review. While this study followed an analytic framework, we did not attempt to present a view regarding the weight of evidence. The results therefore did not seek to assess the quality of evidence and cannot determine if the findings are generalizable. Having charted the data, however, we were able to present our findings in a narrative format after organising the data according to study characteristics (Table 3) and themes (Table 4).

**Table 3.** Study characteristics of included data.

| Author | Study type |
| --- | --- |
| [17–20] | Randomised control trials (RCT) |
| [21] | Meta-analysis of randomised control trials |
| [22] | Clinical review |
| [4], [7], [23–24] | Systematic review and meta-analysis |
| [3], [5–6], [25–29], [63] | Systematic review |
| [30] | Mixed methods |
| [31–34] | Literature review / scoping review |
| [36–37] | Integrative review |
| [38–50] | Pilots / trials |
| Author | Demographics |
| [35], [42–43], [47], [53], [58], [59] | Young people aged 14–22 years old |
| [62] | Youth |
| [54] | Young females |
| [12], [23], [28], [45], [49], [55] | Young people, adults, carers, parents and/or healthcare professionals |
| [5], [38], [40], [51], [59] | Adults aged 24-61 years old |
| [37], [46] | Adult males |
| [48] | Older adults aged over 50 years old |
| [20], [28], [56], [60] | Veterans |
| [26], [39], [57], [59], [64] | Ethnicity focus on Indigenous cultures |
| [21], [25], [27], [29], [30-34], [36], [38], [50-52], [65-72] | Not specified or implied all populations were included |

**Table 4.**
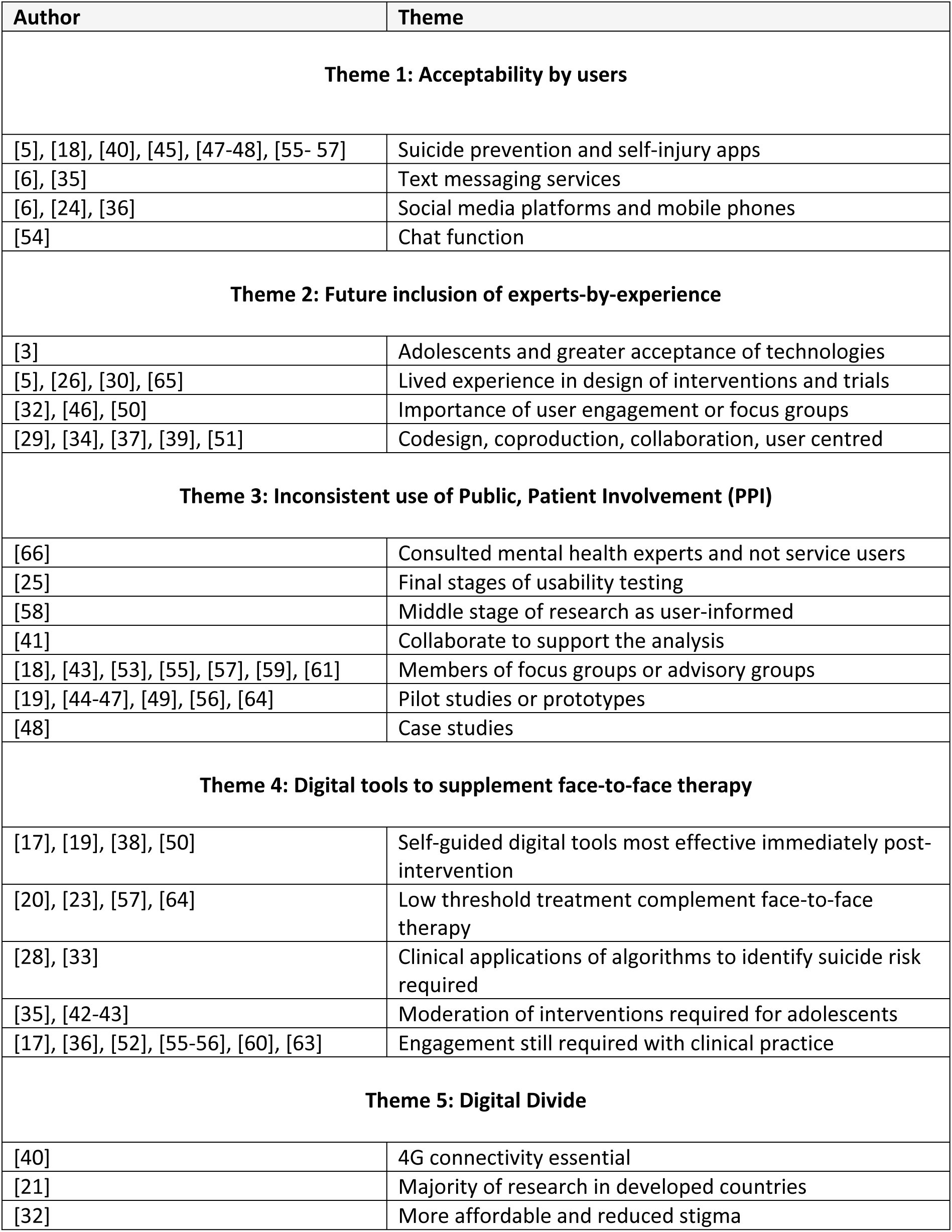
Narrative grouping of data into themes.

| Author | Theme |
| --- | --- |
| **Theme 1: Acceptability by users** |  |
| [5], [18], [40], [45], [47-48], [55- 57] | Suicide prevention and self-injury apps |
| [6], [35] | Text messaging services |
| [6], [24], [36] | Social media platforms and mobile phones |
| [54] | Chat function |
| **Theme 2: Future inclusion of experts-by-experience** |  |
| [3] | Adolescents and greater acceptance of technologies |
| [5], [26], [30], [65] | Lived experience in design of interventions and trials |
| [32], [46], [50] | Importance of user engagement or focus groups |
| [29], [34], [37], [39], [51] | Codesign, coproduction, collaboration, user centred |
| **Theme 3: Inconsistent use of Public, Patient Involvement (PPI)** |  |
| [66] | Consulted mental health experts and not service users |
| [25] | Final stages of usability testing |
| [58] | Middle stage of research as user-informed |
| [41] | Collaborate to support the analysis |
| [18], [43], [53], [55], [57], [59], [61] | Members of focus groups or advisory groups |
| [19], [44-47], [49], [56], [64] | Pilot studies or prototypes |
| [48] | Case studies |
| **Theme 4: Digital tools to supplement face-to-face therapy** |  |
| [17], [19], [38], [50] | Self-guided digital tools most effective immediately post-intervention |
| [20], [23], [57], [64] | Low threshold treatment complement face-to-face therapy |
| [28], [33] | Clinical applications of algorithms to identify suicide risk required |
| [35], [42-43] | Moderation of interventions required for adolescents |
| [17], [36], [52], [55-56], [60], [63] | Engagement still required with clinical practice |
| **Theme 5: Digital Divide** |  |
| [40] | 4G connectivity essential |
| [21] | Majority of research in developed countries |
| [32] | More affordable and reduced stigma |

DW reviewed the first iteration of themes according to the inclusion criteria and analysed iteratively the emerging categories from the published and grey literature. MN analysed the emerging themes and referred to the full text where required. DW and MN worked together to simplify the descriptions and identified fuller commonalities according to the study characteristics and five themes. A final review was provided by the rest of the research team to confirm the results.

## Results

### Study characteristics of the included studies

Table 3 provides an overview of the study characteristics. There were several study types from the 52 published articles including four randomized control trials [17–20], one meta-analysis of randomised control trials [21], one clinical review [22], four systematic review and meta-analysis [4, 7, 23, 24], nine systematic reviews [3, 5, 6, 25–29, 63], one mixed methods [30], five literature or scoping reviews [31–35], two integrative reviews [36–37], thirteen pilots or trials [38–50], seven qualitative studies [51-57), two sequential cross-sectional mixed methods (58–59) and two proof-of-concept [60–61].

The following demographics were noted from the published and grey literature. 7 x articles identified young people specifically ranging from 14-22 years old [35, 42–43, 47, 53, 58–59). 1 x website targeted youth [62] and 1 x article focused on young females [54]. 6 x articles focused on young people alongside adults, carers, parents and/or healthcare professionals [12, 23, 28, 45, 49, 55]. Adults were identified within 6 articles with an age range from 24-61 years [5, 38, 40, 51, 59]. Two articles specifically discussed adult males [37, 46].

Older adults aged over 50 years were discussed in one article [48]. Veterans were discussed in four articles [20, 28, 56, 60]. Ethnicity was focused on the experiences of Indigenous cultures within 5 articles from Australia, New Zealand, Canada and USA [26, 39, 57, 59, 64]. 16 published articles and 8 items of grey literature did not specify which populations the authors were targeting, or it was implied that all populations were included [21, 25, 27, 29–34, 36, 38, 50–52, 65–72].

### Theme One: Acceptability by users

Table 4 provides an overview of the five themes that emerged from the 52 peer reviewed articles and 9 items from grey literature. We found the first theme of *acceptability by users* was prominent from the published literature (n=19). The articles either examined acceptability of digital tools for suicide prevention explicitly or it emerged as a key finding [5–6, 18, 23–24, 35–40, 45, 47–49, 54–57]. Interestingly, almost half of the 19 items (n=9) specifically reviewed suicide prevention and self-injury apps to establish if users found them acceptable [5, 18, 40, 45, 47–48, 55–57]. The remaining published literature focused on text messaging services [6, 35] or involved systematic reviews of social media platforms and mobile phone-based interventions [6, 24, 36]. Only one article reviewed the chat function on the Samaritan’s website, a major non-government organisation that has a long history of providing suicide prevention services [54].

### Theme Two: Future inclusion of experts-by-experience

The second theme identified from 12 articles located in the published literature recommended the *future inclusion of experts-by-experience* [3, 5, 26, 29, 30, 32, 34, 37, 39, 46, 50, 51]. One item from grey literature also emerged within this theme [65]. The terminology and ways to involve experts-by-experience varied among the literature. For example, involving people with *lived experience* in the design of interventions and trials was specifically highlighted in four studies [5, 26, 30, 65]. *Codesign, coproduction, user-centred design and collaboration* were terms used in five studies where the emphasis was on end-to-end input, albeit in a generalised format [29, 34, 37, 39, 51]. To a lesser extent, the importance of *user engagement* or *focus groups* was recommended in three studies [32, 46, 50] and one study specifically recommended targeting *adolescents* whom the authors argued have a greater acceptance of technologies [3]. Interestingly, given that the recommendations from these articles were to include experts-by-experience within health research, we found it surprising that such recommendations were identified and not followed through by the authors themselves. We found one exception where the Samaritans’ website was reviewed from 2017-2019 [65]. The review involved repackaging the Samaritans’ web-based services and text service, SHOUT. Following consultation with people with lived-experience a new website (StayingSafe) included strategies for developing a safety plan and lived-experience video footage. The review found that it was important to continue evaluating developments and identify models of effective help provision. In addition, it was noted that users should be involved not only in such evaluation, but also as co-producers of new online materials and approaches.

### Theme Three: Inconsistent use of Patient and Public Involvement (PPI)

Patient and Public Involvement (PPI) in research is defined as ‘Research being carried out *with* or *by* members of the public rather than *to, about* or *for* them’ [73]. Notwithstanding, *the Inconsistent use of PPI theme* emerged from 19 published articles where the authors where unclear about the purpose and role of PPI [18–19, 25, 41, 43–49, 53, 55–58, 61, 64]. Furthermore, Facebook, a major online networking site, stated that their company liaised with mental health experts (not service users) and considered this type of consultation met the requirements of PPI research [66]. None of the data from the published and grey literature identified PPI as co-authors and used a variation of terms to imply codesign methodology. For example, PPI were involved passively during the final stages of usability testing for digital tools [25], the middle of the research as user-informed participants [58], part of a collaborative process to support the analysis [41], as members of focus groups or advisory groups [18, 43, 53, 55, 57, 59, 61], pilot studies or prototypes [19, 44–47, 49, 56, 64] and case studies [48].

### Theme Four: Digital technology tools to supplement face-to-face therapy

The theme of *digital technology tools as a supplement to face-to-face therapy*, emerged from 20 articles within the published literature [7, 17, 19, 20, 23, 28, 33, 35–36, 38, 42–43, 50, 52, 55–57, 60, 63–64]. From within this theme, four articles found that self-guided digital tools were most effective immediately post-intervention [17, 19, 38, 50]. It was not clear however if the effectiveness continued once the studies were complete and the digital tools such as tablets and mobile phones were returned to the researchers. Four articles noted that low threshold treatments were successful as a complement face-to-face therapy [20, 23, 57, 64]. People that were considered actively at high risk of suicide were not included in the studies. Websites and platforms such as Facebook were reviewed where clinical applications of algorithms were recommended as a future requirement to identify suicide risk [28, 33]. As a high risk group of suicide, moderation of digital interventions were recommended in three studies particularly for adolescents [35, 42–43] and engagement with clinical practice is still required [17, 36, 52, 55–56, 60, 63]. Although the articles recommend digital intervention as part of model of care, 9 articles involved limited sample sizes and were not generalizable [38, 60, 23, 52, 56, 20, 57, 64, 7].

### Theme Five: Digital Divide

The digital divide is the gap that exists between individuals who have access to modern information and communication technology and those who lack access [74]. Although only three articles were included within the theme of *digital divide*, we felt its impact required attention as disparities associated with access, geography and income were identified [21, 32, 40].

One article noted that 4G connectivity was essential for reliable and continuous access to the applications [40]. Those on low incomes tended to use 2G and 3G technology that did not require smartphones and relied on text messaging services. Similarly, a second article found that technology-enhanced interventions such as telephone management were more affordable than face-to-face interventions and reduced stigma [32]. Furthermore the majority of research on digital technology and suicide prevent was conducted in First-world countries, which excluded less developed countries [21]. Recommendations include policy makers improving peoples’ digital literacy and inclusion of research with more countries on a global scale [21].

### Stage Six: Stakeholder consultation

The experts-by-experience members of this project are part of the university’s PPI group and regularly provide input into programmes or research. The group were approached by the primary researcher to be part of the project and were paid as per the National Institute of Health and Care Research guidelines [73]. The members met on a monthly basis to review the articles and the chairperson meet with the primary researcher in between meetings. All members have been identified on funding applications, publications and conference presentations.

## Discussion

This scoping review was the first to explore what is known about codesign, digital tools and suicide prevention. Our findings highlighted the disconnect between developers, researchers and experts-by-experience understanding and acceptability of the digital health tools. For example, none of the published articles were clear about codesign as a methodology and end users were only included passively through focus groups, advisory groups, or during the testing stage of pilot studies or prototypes. Despite the initial engagement of experts-by-experience with digital tools for suicide prevention immediately after an intervention, the removal of regular monitoring by health professionals once trials were completed, contributed to lack of data to confirm ongoing success.

Supplementation of digital solutions in partnership with face-to-face therapy will increase especially for digital natives such as adolescents. The ethical concerns over how younger populations will have their privacy safeguarded is an area that is yet to be developed more fully within the literature. The digital divide is noteworthy as a future concern as countries such as the United Kingdom are phasing out 2G and 3G networks in 2023 to make room for faster 5G networks [75]. The implications for those on low incomes and experiencing mental distress may not be visible to the general public as access to digital platforms in the future will require uninterrupted connectivity through smartphones and internet access.

## Limitation

A limitation of this scoping review is the exclusion of literature that was not written in the English language. Based on feedback on our scoping protocol, we extended our search to include countries from the European Union and countries such as China, Japan and Sri Lanka. The search criteria would be extended to non-English articles for future reviews.

## Conclusion

When we conducted this scoping review from a codesign perspective, we were surprised by the lack of literature published and unpublished, that also followed the same methodology. We believe our findings will support future researchers and developers to implement codesign that maintains the integrity of their project. Authentic involvement requires experts-by-experience as co-authors and end-to-end partners from design, implementation and evaluation of digital health tools for suicide prevention.

## Data Availability

No data sets were generated or analysed during the current study. All relevant data from this study will be made available upon completion of the study.

## Supporting information

S1 Table. Population, Concept, Context Search String.

(DOCX)

S2 Table. Data Extraction.

(DOCX)

S3 Table. Study characteristics of included data.

(DOCX)

S4 Table. Narrative grouping of data into themes.

(DOCX)

S1 Figure. PRISMA flow diagram for included searches of databases and grey literature.

(DOCX)

## Acknowledgments

We acknowledge the support and guidance from Simon Couth, The Working Academy, Academic Librarians and members of the expert-by-experience group.

## Author Contributions

Conceptualisation: Dianne Wepa

Investigation: Dianne Wepa, Martin Neale, Waseem Abo-Gazala, Sally Cusworth, Jae Hargan, Manoj Mistry, Jimmy Vaughan, Stephen Giles, Mehnaz Khan

Methodology: Dianne Wepa, Martin Neale, Waseem Abo-Gazala

Formal analysis: Dianne Wepa, Martin Neale

Writing – original draft: Dianne Wepa

Writing – reviewing & editing: Dianne Wepa, Martin Neale, Waseem Abo-Gazala, Sally Cusworth, Jae Hargan, Manoj Mistry, Jimmy Vaughan, Stephen Giles, Mehnaz Khan

